# "Correlation Between Clinical Presentation and Brain CT Findings in Acute Dizziness: A Retrospective Cross-Sectional Analysis at a Tertiary Referral Center"

**DOI:** 10.64898/2026.06.25.26356549

**Authors:** Alireza Abbasi, Maryam Moghbel Baerz, Mohammad Farhadi, Robab Sadegh, Kiarash Kavari, Faezeh Rastaghi, Zeinab Azadian, Amir Hossein Rajabi, Amin Nasr

## Abstract

**Background:** Dizziness is a frequent presenting complaint in the emergency department (ED), prompting extensive diagnostic evaluation. Non-contrast brain computed tomography (CT) is often utilized to rule out serious central pathologies, but its diagnostic yield is debated, leading to concerns about overuse. This study aimed to identify clinical predictors associated with abnormal brain CT findings in patients with acute dizziness to help refine imaging selection criteria.

**Methods:** We conducted a retrospective analysis of 291 consecutive adult patients who presented with new-onset dizziness and underwent a non-contrast brain CT scan at Namazi Hospital, a tertiary referral center, between January 2019 and 2021. Patient data, including demographics, comorbidities, clinical symptoms, and hospital outcomes, were extracted from medical records. Statistical analyses were performed to determine associations between clinical variables and CT findings, with odds ratios (OR) and 95% confidence intervals (CI) calculated.

**Results:** The diagnostic yield of brain CT was low, with a significant majority of scans (72.2%, n=210) revealing no acute pathology. Key clinical factors predicting abnormal CT findings included a history of diabetes mellitus, the presence of ataxic gait, and headache. Conversely, nausea and vomiting were significant predictors of normal findings, being associated with lower odds of central pathology.

**Conclusion:** The diagnostic yield of routine brain CT in patients with acute dizziness is low. However, specific clinical indicators can effectively stratify risk. The presence of focal neurological signs like ataxia, headache, and certain comorbidities such as diabetes should heighten suspicion for central pathology and support the use of neuroimaging. In contrast, isolated vestibular symptoms like nausea and vomiting are associated with a lower probability of abnormal findings. These results could inform the development of clinical decision rules to optimize CT utilization, thereby reducing unnecessary radiation exposure and healthcare costs.

**Key Points:**

- A large proportion of dizzy patients undergoing non-contrast head CT in the emergency department have no acute radiological abnormalities, underscoring the low overall diagnostic yield of routine CT in this setting.
- In our cohort, diabetes mellitus, ataxic gait, and headache were independently associated with abnormal CT findings, suggesting that these clinical features may help identify patients at higher risk for central pathology.
- Nausea and vomiting were more commonly observed in patients with normal CT scans, consistent with their frequent association with peripheral vestibular syndromes rather than central causes.
- Incorporating simple bedside findings, such as gait assessment and the presence or absence of headache into imaging decisions may help reduce low-yield CT utilization while maintaining attention to high-risk presentations.
- Prospective, multicenter validation of clinically anchored imaging decision rules is needed before these findings can be translated into definitive practice recommendations.

## Background

Dizziness is a non-specific term encompassing various sensations of disturbed spatial orientation, including vertigo, unsteadiness, and lightheadedness (Bárány Society, ICVD) (1). Vertigo is specifically defined as the sensation of self-motion when no self-motion is occurring, or an exaggerated sense of motion during normal head movement (2).

Dizziness arises from causes ranging from benign vestibular neuritis to life-threatening stroke or arrhythmia (3). Approximately 15% to 20% of adults experience dizziness, including vertigo, each year, according to large population-based studies. It affects roughly two to three times as many women as men, and its prevalence increases with age (4).

A review of emergency department visits in the United States between 1995 and 2004 found that vertigo and dizziness together accounted for only 2–5% of all presentations (5).

Compared to patients with other primary complaints, these patients have higher rates of resource use and length of stay in the emergency department (5, 6).

The primary etiological diagnosis of vertigo and dizziness relies on a classification scheme that considers both the nature of the disease and its anatomical origin. However, accurately localizing and diagnosing vestibular disorders including peripheral, central, psychiatric, and functional etiologies can be difficult in some patients (7, 8).

Most acutely dizzy patients in the emergency department (ED) present in one of three patterns according to a timing and triggers paradigm: acute vestibular syndrome (AVS), spontaneous episodic vestibular syndrome (s-EVS), and triggered episodic vestibular syndrome (t-EVS).

These categories inform the differential diagnosis, diagnostic testing, and interpretation of many of these tests (9).

The most frequent cause of dizziness is a peripheral etiology. Issues with the inner ear or the vestibular nerve, which links the inner ear and brain, can result in peripheral vertigo (10). The most frequent peripheral causes of dizziness are vestibular neuritis, Ménière’s disease, and benign paroxysmal positional vertigo (3).

Given non-contrast CT’s extremely low sensitivity for acute ischemic stroke and the correspondingly low pretest probability of stroke among unselected patients with acute dizziness, routine head CT provides limited diagnostic value in this population (11, 12). Even though intracerebral hemorrhage (ICH) is much more sensitive to CT scans, ICH is a rare cause of isolated dizziness in patients (13).

CT might make sense if the dizziness is not isolated or is accompanied by a severe headache. In that situation, a positive CT scan is helpful, but a negative one shouldn’t be comforting (9).

Only 3–6% of ED patients with acute dizziness had major central causes, primarily ischemic stroke, according to studies (14). The percentage of ED patients who are discharged with a peripheral vestibular diagnosis and later readmitted with an acute stroke is extremely low, ranging from 0.14 percent to 0.5 percent, due to the fact that peripheral causes of acute dizziness in the ED are far more common than central ones, and many patients whose cause is a stroke are correctly diagnosed (15, 16).

In the emergency department, CT is used much more often than MRI to try to diagnose or rule out stroke, but the results are rarely useful. Emergency clinicians have a significant knowledge gap in this area (17, 18). Also overuse and almost routine CT scans in the emergency department can have a number of negative consequences for patients and hospitals, such as needless radiation exposure and lengthening hospital stays (19).

We conducted this retrospective study based on the hypothesis that certain clinical features, particularly focal neurological deficits and specific comorbidities, would be significant predictors of abnormal CT findings in patients with new-onset dizziness. The primary aim was to identify these predictors and explore the feasibility of developing a simple decision rule to optimize the use of neuroimaging in the emergency department.

## Methods

### Study Design and Setting

We conducted a retrospective, cross-sectional analysis of patient records at Namazi Hospital, a tertiary referral center in Shiraz, Iran. The study included patients who presented to the Neurology Department between January 2019 and December 2021. Ethical approval was obtained from the Shiraz University of Medical Sciences Ethics Committee (IR.SUMS.MED.REC.1401.467), and the study was conducted in accordance with the principles of the Helsinki Declaration.

### Study Population

The target population consisted of consecutive adult patients (≥18 years old) who presented with new-onset dizziness and underwent a non-contrast brain CT scan as part of their initial evaluation. Patients were excluded if they had: (1) incomplete or missing medical records; (2) a known, active pre-existing intracranial pathology directly explaining the symptoms; (3) recent head trauma (<6 months); or (4) dizziness definitively attributed to non-neurological or isolated peripheral causes at the initial emergency presentation (e.g., orthostatic hypotension, a confirmed peripheral vestibular condition such as BPPV, or metabolic disruption) that obviated the need for neuroimaging.

However, it is important to clarify that patients presenting with acute dizziness who had a past medical history of central nervous system pathologies (e.g., a prior resolved stroke or a previously treated brain tumor) were included. The rationale for their inclusion is that such a medical history places these patients at a significantly higher baseline risk for acute central neuro-otologic events, thereby justifying the clinical decision to perform a brain CT scan.

### Variables and Data Collection

Data were extracted from archived medical records using a standardized case report form [see Additional file 1]. The collected variables included:

Demographics: Age and sex, Clinical Data: Comorbidities (e.g., diabetes mellitus, hypertension, valvular heart disease), current medications, and presenting symptoms (e.g., headache, nausea, vomiting, ataxic gait), Radiological Findings: Brain CT results were categorized as either normal (no acute pathology) or abnormal (e.g., infarction, intracranial hemorrhage, neoplasm), Hospital Outcome: Final disposition was recorded as discharged, admitted, or died.

### CT Imaging Protocol

All non-contrast brain CT scans were performed on a 128-slice GE Revolution scanner using a standardized protocol (120kVp; 250mAs; slice thickness <5mm for posterior fossa evaluation). To ensure objectivity, scans were independently reviewed by two radiology residents who were blinded to the clinical data. Discrepancies were adjudicated by a board-certified neuroradiologist with seven years of experience.

### Statistical Analysis

All statistical analyses were performed using SPSS version 25, to account for multiple comparisons in our analyses, we applied the False Discovery Rate (FDR) control method. Consequently, adjusted p-values, reported as q-values, were calculated. Statistical significance was set at q < 0.05.. Continuous variables (e.g., age) were tested for normality and reported as mean ± standard deviation; group differences were assessed using an independent-samples t-test or Mann-Whitney U test. Categorical variables were reported as counts and percentages, and associations with CT outcomes were evaluated using the Pearson chi-square or Fisher’s exact test. Odds ratios (OR) with 95% confidence intervals (CI) were calculated to quantify the strength of association between clinical predictors and abnormal CT findings.

While baseline clinical variables were assessed as independent predictors of abnormal CT findings, the final disposition (hospital admission or death) was also included in the multivariable model. This was done not to treat disposition as a preceding predictor, but rather to quantify the independent strength of the association between positive neuroimaging findings and adverse clinical trajectories, adjusting for baseline confounders.

## Results

The study included 291 patients with a mean age of 59.4 ± 18.7 years; 146 (50.2%) were male. The most prevalent comorbidities were hypertension (29.2%), diabetes mellitus (15.1%), and coronary artery disease (7.6%). The most frequent presenting symptoms were nausea (34.4%), vomiting (31.6%), and headache (29.6%). (Table 1)

**Table 1.** Baseline Demographic and Clinical Characteristics of Patients (n=291)

| Characteristic | n (%) |
| --- | --- |
| **Demographics** |  |
| Age (years), Mean ± SD | 59.4 ± 18.7 |
| Sex, Male | 146 (50.2) |
| **Past Medical History** |  |
| Hypertension | 85 (29.2) |
| Diabetes Mellitus | 44 (15.1) |
| Coronary Artery Disease | 22 (7.6) |
| Malignancy | 10 (3.4) |
| Chronic Kidney Disease | 9 (3.1) |
| Myocardial Infarction | 9 (3.1) |
| Valvular Heart Disease | 4 (1.4) |
| **Presenting Symptoms** |  |
| Nausea | 100 (34.4) |
| Vomiting | 92 (31.6) |
| Headache | 86 (29.6) |
| Ataxic Gait | 36 (12.4) |
| Positional Symptoms | 28 (9.6) |
| Unilateral Weakness | 16 (5.5) |
| Double Vision | 16 (5.5) |

A significant majority of patients (72.2%, n=210) had normal brain CT scans. Among the 81 (27.8%) abnormal scans, the most common findings were cerebral infarction (12.0% of the total cohort) and neoplasm (10.3%). The final hospital disposition was discharge for 56.4% of patients, while 1.4% died. (Table 2)

**Table 2.** CT Scan Findings and Hospital Disposition (n=291)

| Outcome | Category |
| --- | --- |
| **CT scan Finding** |  |
| Normal | 210 (72.2) |
| Abnormal | 81 (27.8) |
| **Type of Abnormal Finding\*** |  |
| Cerebral Infarction | 35 (12.0) |
| Neoplasm | 30 (10.3) |
| Intracranial Hemorrhage | 9 (3.1) |
| Infection | 4 (1.4) |
| Other | 3 (1.0) |
| **Hospital Disposition** |  |
| Discharged | 164 (56.4) |
| Admitted | 112 (38.5) |
| Transferred | 11 (3.8) |
| Died | 4 (1.4) |

### Primary Predictors of Abnormal CT Findings

Several clinical factors were independently associated with abnormal CT findings. Regarding clinical trajectories, an abnormal CT finding was strongly and independently associated with a higher likelihood of hospital admission or death; patients who were admitted or died had eight times the odds of an abnormal scan compared to those discharged (OR 8.01, 95% CI [4.25–15.10], *q* < 0.001). Significant comorbidities predicting abnormal findings included valvular heart disease (OR 4.75, *q* = 0.049) and diabetes mellitus (OR 3.10, *q* = 0.004). The presence of ataxic gait (OR 2.41, *q* = 0.028) and headache (OR 1.83, *q* = 0.049) also predicted abnormal scans. Conversely, nausea (OR 0.45, *q* = 0.028) and vomiting (OR 0.48, *q* = 0.028) were significantly associated with a lower likelihood of abnormal CT results, acting as protective factors. (Table 3)

**Table 3.** Multivariable Logistic Regression Analysis of Clinical Predictors and Associated Outcomes for Abnormal Brain CT Findings.

| Variable | Comparison | Odds Ratio (95% CI) | Adjusted *q*-value |
| --- | --- | --- | --- |
| **Patient History** |  |  |  |
| Diabetes Mellitus | Present vs. Absent | 3.10 (1.52 – 6.33) | 0.004 |
| Valvular Heart Disease | Present vs. Absent | 4.75 (1.02 – 22.10) | 0.049 |
| No Prior Medical History | Yes vs. No | 0.42 (0.25 – 0.70) | < 0.001 |
| **Clinical Findings** |  |  |  |
| Headache | Yes vs. No | 1.83 (1.03 – 3.25) | 0.049 |
| Nausea | Yes vs. No | 0.45 (0.24 – 0.84) | 0.028 |
| Vomiting | Yes vs. No | 0.48 (0.26 – 0.88) | 0.028 |
| Ataxic Gait | Present vs. Absent | 2.41 (1.18 – 4.93) | 0.028 |
| **Associated Clinical Outcome** |  |  |  |
| Final Disposition | Admitted/Died vs.<br>Discharged | 8.01 (4.25 – 15.10) | < 0.001 |

CI, Confidence Interval. Adjusted *q*-values are corrected for multiple comparisons using the false discovery rate method. Statistically significant values ( $q < 0.05$ ) are shown in bold

### Clinically Relevant Subgroup Analyses

In patients younger than 60, nausea (OR 0.15, *q* = 0.010) and vomiting (OR 0.20, *q* = 0.017) were strong predictors of normal CT scans, while final disposition remained the most associated clinical outcome of abnormal findings (OR 37.9, *q* = 0.004). In contrast, for patients over 60, none of the individual clinical symptoms were significant, and only final disposition had a significant association (OR 6.40, *q* < 0.001).

Analysis by sex revealed that nausea was a significant predictor of normal CT findings in women (OR 0.30, *q* = 0.02), while no individual symptom was predictive in men. For men, only the final hospital disposition was significantly associated with abnormal CT results (OR 7.64, *q* = 0.008). (Table 4)

**Table 4.** Subgroup Analysis of Key Predictors for Abnormal Brain CT Findings.

|  | Male | Female | Age < 60 | Age ≥ 60 | Hypertension | Diabetes | Antiplatelet Users | Overall Cohort |
| --- | --- | --- | --- | --- | --- | --- | --- | --- |
| Predictor | OR (95% CI) | OR (95% CI) | OR (95% CI) | OR (95% CI) | OR (95% CI) | OR (95% CI) | OR (95% CI) | OR (95% CI) |
| Headache | 0.67(0.31–1.45) | 0.94(0.42–2.10) | 1.54(0.63–3.77) | 1.17(0.58–2.34) | 1.00(0.34–2.94) | 1.31(0.26–6.59) | 1.05(0.38–2.91) | 1.51 (0.64–3.56) |
| Nausea | 0.67(0.30–1.50) | 0.30(0.12–0.77) * | 0.15(0.04–0.56) * | 0.76(0.38–1.52) | 2.38(0.6–8.86) | 0.29(0.03–2.67) | 0.77(0.31–1.93) | 0.69 (0.28–1.66) |
| Vomiting | 0.57(0.25–1.28) | 0.40(0.16–1.01) | 0.20(0.05–0.76) * | 0.72(0.36–1.45) | 2.25(0.6–7.85) | 0.24(0.03–2.22) | 0.73(0.28–1.85) | 0.70 (0.29–1.71) |

OR, Odds Ratio; CI, Confidence Interval
\* $q < 0.05$ , statistically significant after correction for multiple comparisons.

Analyses of small, underpowered subgroups, such as patients on anticoagulant therapy (n=9) or with coronary artery disease (n=22), were found to be inconclusive.

## Discussion

The principal finding of this study is that while the overall diagnostic yield of non-contrast brain CT in unselected emergency department patients with acute dizziness is low, specific clinical features can effectively stratify patients into high- and low-risk groups for serious central pathology. We identified focal neurological deficits, specifically ataxic gait, and a history of diabetes mellitus as independent predictors of abnormal CT findings. Conversely, the presence of classic vestibular symptoms, such as nausea and vomiting, was associated with normal imaging, suggesting a peripheral etiology.

Our finding that headache is an independent predictor of abnormal neuroimaging suggests its potential value for refining clinical risk-stratification models. This indicates that headache warrants consideration as a key variable in the development of future decision tools for dizzy patients. Its presence could help clinicians better identify a subset of patients who may benefit from a lower threshold for CT scanning, thereby potentially improving the diagnostic yield of the evaluation process There have been conflicting findings from earlier research on the diagnosis of clinically significant findings in head CT scans in patients experiencing vertigo.

Our overall yield of positive CT findings (27.8%) exceeds that reported in several prior series. Prior findings regarding neuroimaging yield in dizziness patients vary significantly based on patient selection and imaging modality.

Pavlović et al discovered that only 6–9% of 72 patients with non-focal dizziness had clinically significant CT abnormalities, and 1.14% of patients had an ischemic lesion (20). By contrast, Ahsan et al. examined patients who experienced vertigo and dizziness with CT or MR, they found 6.17% positive results and 0.74% clinically significant results; they excluded patients who had a history of stroke, brain tumor, brain surgery, or other neurological disorders (18).

The broader inclusion criteria in our study, which allowed for patients with pre-existing conditions such as prior stroke or tumors, may have introduced a referral bias toward more complex clinical presentations.

In another study of the patients who underwent neuroimaging (CT and MRI) in the emergency department, 13 percent (n = 42) were eventually diagnosed with serious neurological disease. Six percent (95 percent CI, 3 percent-9 percent) of CT scans detected a relevant abnormal finding, while nine percent (95 percent CI, 4 percent-15 percent; P = 0.31) of MRIs did the same. Nearly all relevant findings were explained by intracerebral hemorrhage (20 percent), neoplasm (24 percent), and infarction (52%) (21).

In a study of 79 dizzy patients, 17% of CT scans and 25% of MRIs revealed an etiological abnormality (when atrophy was removed as a relevant finding) (22).

There were no acute abnormalities on CT imaging in a prospective study of 200 patients who had acute vertigo or dizziness as their primary complaint in the emergency department. However focal neurological deficits, headaches, or trauma patients were excluded (23).

In a three-year study, brain CT scans were performed on 48% of ED patients who experienced vertigo and dizziness. Out of these, only 0.74% produced pathology that was clinically significant (24).

A number of factors, including different scanning protocols, broader inclusion criteria (we did not exclude patients with non-focal neurologic symptoms or vascular risk factors), and possible referral bias due to more complex or higher-risk patients being preferentially imaged, could account for our significantly higher positive CT rate (27.8%) compared to about 6% in other CT-only series (18).Additionally, older age (mean age ∼59 vs. ∼54 in Pavlović) and higher prevalence of cerebrovascular risk factors may increase stroke probability (20).

Failure to address these points risks overestimating CT utility in dizziness protocols. Among associated clinical symptoms, nausea (34.4%) and vomiting (31.6%) were the most frequent. Vertigo and dizziness typically accompany autonomic symptoms like nausea and vomiting as well as unsteadiness (3).

There is evidence that vertigo is more common and severe in women than in men, with higher prevalence rates, higher levels of disability, and a stronger correlation with anxiety and depression seen in studies (25). About half of the dizziness patients in this study were men, with the remaining patients being women.

Age-stratified analysis revealed distinct clinical predictors: nausea was significantly associated with normal CT findings in patients under 60 years, whereas headache was a significant predictor of abnormal findings in those over 60, the former reported noticeably more complaints of true vertigo and related nausea and vomiting. Elderly patients frequently reported experiencing unsteadiness or falling (26).

different pattern of association between clinical symptoms and radiological findings was seen in the two age groups.

The two most common underlying diseases in the current study were high blood pressure (29%) and diabetes mellitus (15%). In a study of 493 elderly subjects, dizziness was significantly associated with hypertension (χ2 = 6.26, p = 0.01) (27). Additionally, patients with diabetes were more likely to experience more severe vertigo symptoms and worse sleep quality (28).

Cerebral infarction (12%) was the most common radiological finding, and given the low diagnostic sensitivity of CT scan in our study in diagnosing acute ischemic stroke, the actual prevalence of infarction is expected to be much higher (29).

While our multivariable model focused on baseline clinical features as pre-test predictors, it also revealed a highly significant, independent association between abnormal CT findings and adverse final dispositions (hospital admission or death). This strong correlation reflects the expected clinical cascade: the detection of acute central nervous system pathologies on neuroimaging directly dictates the need for urgent inpatient management and inherently correlates with a higher risk of adverse outcomes. For context, the in-hospital mortality rate in our study was 1.4%, whereas a previous study reported a 30-day mortality rate of 0% and a 100-day mortality rate of 1.0% (2/207) for patients presenting with dizziness as their primary complaint (30).

Our study demonstrated a negative association between nausea/vomiting and abnormal CT findings. This should not be interpreted as a biological protective effect, but rather as a clinical signifier pointing toward a peripheral etiology. Nausea and vomiting are autonomic symptoms of acute peripheral vestibulopathies, such as vestibular neuritis (31). As these conditions originate from the inner ear labyrinth and are anatomically distinct from the brain parenchyma, they yield normal findings on non-contrast CT scans.

The most common cause of severe central vestibular dysfunction is an ischemic stroke of the posterior fossa, which requires prompt recognition for potential emergent treatment like intravenous thrombolysis (32). For patients presenting with an Acute Vestibular Syndrome (AVS) our results could provide empirical evidence to help differentiate its cause. The presence of prominent nausea and vomiting, in the absence of other focal neurological deficits, suggests a peripheral AVS. This aligns with a low pre-test probability for central pathology, representing a low-yield population for routine neuroimaging. Conversely, the presence of findings such as ataxic gait in a dizzy patient even with less severe vomiting should heighten suspicion for a central AVS, such as a posterior circulation stroke involving the cerebellum or brainstem, and thus warrants immediate imaging (33).

The diagnostic accuracy of non-contrast CT in determining the primary causes of acute dizziness, particularly stroke, is notoriously low. Consequently, it frequently fails to detect acute ischemic strokes in the early phases of symptom onset. Non-contrast CT is still reasonably accurate in detecting acute brain hemorrhages, despite its poor diagnostic precision in ischemic stroke detection. As a result, it might still be the first option (24).

## Limitations

Our study has a number of significant limitations. Its retrospective, single-center design, which depends on varying clinical documentation, is prone to selection bias. Additionally, our results might not apply to community settings because we only examined non-contrast head CT scans at a tertiary academic emergency department. Third, the actual prevalence of acute ischemic strokes and other central pathologies was most likely underestimated because we did not conduct any MRI (or other follow-up imaging) on patients who had normal CT findings. Fourth, although the first 24 hours may see radiographically occult early ischemic changes, the time interval between the onset of dizziness and CT acquisition was not documented. Lastly, we did not follow up with patients after their initial ED visit; as a result, we are unable to determine long-term mortality, extended functional outcomes, or delayed stroke diagnoses.

## Future Directions

Future research should focus on prospective, multicenter studies that enroll a broad spectrum of patients presenting with acute dizziness and vertigo across different levels of care, including community and non-tertiary hospitals. Such studies should incorporate standardized clinical assessment tools, such as structured timing-and-triggers characterization and validated bedside oculomotor examinations (e.g., HINTS-plus protocols), as well as systematic documentation of symptom onset-to-imaging intervals.

In addition, combining non-contrast CT with targeted use of MRI and vascular imaging in predefined risk strata would allow a more accurate estimate of the true prevalence of central pathology and a more precise evaluation of imaging performance. Ultimately, integrating clinical predictors identified in our study, such as diabetes, ataxic gait, and headache with other findings into simple, prospectively validated decision rules may help emergency clinicians safely reduce low-yield CT utilization while maintaining vigilance for central causes of dizziness.

## Conclusion

In this retrospective cohort of adult patients presenting with acute dizziness who underwent non-contrast head CT, we found that diabetes mellitus, ataxic gait, and headache were significantly associated with abnormal CT findings, whereas nausea and vomiting were more frequently observed in patients with normal scans.

These results suggest that brain CT may be most informative in dizzy patients who present with focal gait disturbance or headache, particularly in the presence of vascular risk factors, and less helpful when dizziness is accompanied predominantly by nausea and vomiting in the absence of other concerning features. While our findings should be interpreted cautiously due to the study’s limitations and the low overall sensitivity of CT for acute ischemic stroke, they support a more selective, clinically driven approach to neuroimaging in patients with acute dizziness in the emergency department.

## Data Availability

All data produced in the present study are available upon reasonable request to the authors.

## List of Abbreviations

AVS: Acute Vestibular Syndrome
BPPV: Benign Paroxysmal Positional Vertigo
CI: Confidence Interval
CT: Computed Tomography
ED: Emergency Department
FDR: False Discovery Rate
GRACE-3: Guidelines for Reasonable and Appropriate Care in the Emergency Department 3
HINTS-plus: Head Impulse, Nystagmus, Test of Skew plus
ICVD: International Classification of Vestibular Disorders
ICH: Intracerebral Hemorrhage
MRI: Magnetic Resonance Imaging
OR: Odds Ratio
SD: Standard Deviation
s-EVS: Spontaneous Episodic Vestibular Syndrome
t-EVS: Triggered Episodic Vestibular Syndrome

## Declarations

### Ethics approval and consent to participate

This retrospective study involving human participants and medical record data was approved by the Ethics Committee of Shiraz University of Medical Sciences (IR.SUMS.MED.REC.1401.467). Due to the retrospective nature of the study and the use of anonymized data from existing medical records, the requirement for informed consent was waived by the ethics committee. The study was conducted in full accordance with the principles of the Declaration of Helsinki.

### Consent for publication

Not applicable.

### Availability of data and materials

The data that support the findings of this study are available from the corresponding author upon reasonable request. The datasets consist of de-identified patient information extracted from archived medical records at Namazi Hospital. Due to ethical and institutional restrictions protecting patient privacy, the raw individual-level data are not publicly available. All aggregated data and statistical results are fully presented in the main manuscript (Tables 1–4).

No additional datasets were generated during the study.

### Authors’ contributions

AA and MF conceptualized and designed the study and had primary responsibility for the overall project. AA, MF, and KK supervised the study and contributed substantially to data interpretation. RS and KK coordinated data acquisition and management. RS performed the statistical analysis and contributed to interpretation of the data. FR, MM, ZA, AR, and AN were responsible for data collection and initial data curation. AA, MF, and AN drafted the manuscript, while RS critically revised it for important intellectual content. All authors read and approved the final manuscript.

## Acknowledgements

Not applicable.

## Additional files

File name: Additional file 1

File format: Microsoft Word document (.docx)

Title of data: Case Report Form

Description of data: This file contains the completed case report form that has been filled out for each individual patient included in the study/article.

